# Systematic differences were identified for symptoms of Omicron infected individuals between early and late stage of the COVID-19 pandemic in China

**DOI:** 10.1101/2025.04.10.25325424

**Authors:** Congying Yang, Yi Zhang, Ying Yang, Baibing Mi, Mingwang Shen, Yunpeng Nian, Jinyu Wang, Suixia Cao, Jingchun Liu, Pouria Abolfazli, Hao Huang, Zhongxi Wei, Lixi Liu, Qian Wu, Tianxiao Zhang, Shaobai Zhang

## Abstract

**Aim:** We aimed to investigate the differences in clinical features (specifically, patterns of symptoms) of the COVID-19 patients in the early and late stage of this large scale COVID-19 outbreak in Shaanxi Province in China based on data from a large scale survey.

**Methods:** This study used baseline data from a large survey to investigate the characteristics of the COVID-19 pandemic in Shaanxi province from Dec.2022 to Jan. 2021. A total of 44 sampling clusters (11 villages in rural areas and 33 residential urban areas) were chosen for enrollment of study participants. A self-developed questionnaire was applied to data collection. A total of 14,744 individuals were included, and data from 12,111 valid participants were collected for further analysis.

The Patients infected before Dec. 15th 2022 were classified as early stage infects patients while those infected after Jan. 1st 2023 as late stage infects patients.

**Results:** Information of 2,854 COVID-19 patients, comprising of 1,710 (16.7%) patients infected during the early stage and 1,144 (11.2%) during the late stage, were extracted from the background survey. Most of the COVID-19 typical symptoms (11 out of 12) were found to be more frequent in COVID-19 patients infected during the early stage compared to those infected during late stage. Nine of them showed statistical significance including fever (χ^2^=93.42, *P*<0.001), cough (χ^2^=8.97, *P*=0.003), muscle or body pain (χ^2^=17.57, P<0.001), headache (χ^2^=8.28, *P*=0.004), sore throat (χ^2^=10.10, *P*=0.002), nausea or vomiting (χ^2^=8.40, *P*=0.004), loss of taste or smell (χ^2^=45.05, *P*<0.001) and chills (χ^2^=8.17, *P*=0.004). The significance of most of these symptoms were kept after being adjusted for multiple variables.

**Conclusion:** Individuals infected with the Omicron variant at an earlier stage of a COVID-19 outbreak are more prone to developing certain symptoms compared to those infected at late stage.

## Introduction

On May 5, 2023, the World Health Organization (WHO) chief declared the end of COVID-19 as a Public Health Emergency of International Concern (PHEIC) after more than three years into the pandemic [1]. However, this does not imply the pandemic itself is over. New variants of SARS-Cov-2 are still emerging and might cause new surges in cases and death. The World Health Organization (WHO) has established a review committee tasked with creating enduring, permanent guidelines for countries on the continuous management of COVID-19 [1]. Despite the substantial volume of research papers published in the past three years related to COVID-19, many characteristics of this respiratory infectious disease still remain largely unknown. Hence, further research on COVID-19 is still needed, as it can enhance our preparedness for the next health emergencies.

Among the myriad research topics concerning COVID-19, the academic community has yet to fully explore the symptomatic manifestations of COVID-19 infected patients. Multiple studies have focused on the differences of COVID-19 symptoms in patients infected with different types of SARS-Cov-2 variants [2–5]. Choi *et al.* constructed a retrospective cohort study of COVID-19 patients younger than 18 years of age admitted to five hospitals in Korea and evaluated the clinical characteristics and outcomes of COVID-19-infected children in the Delta variant and the Omicron variant based on a retrospective cohort of COVID-19 patients, and concluded that there were more COVID-19 infected children and complex comorbidities patients in Korea during the Omicron period, compared with the Delta [2]. Justin *et al.* used a self-reporting system to compare the frequency of symptoms in hospital personnel infected with COVID-19 before and after the emergence of the omicron variant, and they concluded that omicron variants were more likely to cause asymptomatic infection [5]. Wang *et al.* analyzed the clinical characteristics of infected patients before and after the emergence of the Omicron variant by conducting a retrospective cohort[6]. It concluded that the syndromes of COVID-19 infected patients during the period of Omicron variant infection were significantly weaker than those during the period of pre-emergence of the Omicron variant and the period of delta variant. Despite of these studies focusing on comparisons of the symptom patterns between patients infected with different types of variants, few reports have been published on whether COVID-19 patients exhibit different symptoms patterns within a single outbreak. By comparing the symptom characteristics of patients at different stages of the same COVID-19 outbreak, factors influencing symptom patterns beyond genetic variations could be explored.

Unlike most other countries around the world, China has adhered to a long-term “dynamic zero-COVID” policy for COVID-19 prevention and control[7–9]. As a result, up until the end of 2022, there have been no large-scale nationwide outbreaks of COVID-19 in the country. On November 11 and December 7, 2022, China announced several new measures to optimize the implementation of COVID-19 epidemic prevention and control [10, 11]. In the subsequent 1-2 months, mainland China experienced a national COVID-19 outbreak [12].

In response to the large-scale COVID-19 outbreak, Xi’an Jiaotong University (XJTU), in collaboration with the Shaanxi Provincial Center for Disease Control and Prevention (SPCDC), initiated a large cohort study to determine the specific modes of COVID-19 transmission and the patterns of symptoms in infected individuals [13].In the present study, we aimed to investigate the differences in clinical features (specifically, patterns of symptoms) of the COVID-19 patients in the early and late stage of this large scale COVID-19 outbreak in Shaanxi Province based on data from this completed survey.

## Materials and Methods

### Study Participants and Data Collection

The present study is based on data from a large-scale COVID-19 survey conducted in Shaanxi Province, China during the COVID-19 pandemic in January 2023 [13]. Shaanxi Province is landlocked, situated in the heart of China, spanning an area of 205,600 km^2^ with a population of 39.2 million, ranking 16th among the provincial-level administrations in China [14]. The study participants were recruited from 44 survey sites across the province, selected through cluster sampling. A total of 14,744 individuals were included, and data from 12,111 valid participants were collected for further analysis.

A self-designed questionnaire was used in this survey, which mainly included the basic information (e.g. sex, age, residence, No. of family members, No. of household transmission cases, pre-existing conditions, basis for diagnosis), the infection and associated symptoms, diagnosis and treatment, and follow-up travel plans of the respondents (A sample of questionnaire and survey protocol were included in the supplementary materials). A face-to-face interview was performed to the study participants to complete the questionnaire, and the questionnaires were distributed online via QQ/We chat platforms if the face-to-face communication was not applicable. Questionnaire data were collected and managed through Research Electronic Data Capture (RED Cap) system[15–17]. Questionnaires with incomplete content, duplicates, or unusual values were judged invalid and were not included in the statistical results. Respondents were fully informed of the purpose and safety of the study before they began to fill out the questionnaire. More specific information of this large-scale survey was recorded in the published article[13].

According to the transmission dynamic data of this pandemic, a plateau shaped peak could be identified from Dec. 15th 2022 to Jan. 1st 2023. Therefore, in the current study we classified those patients infected before Dec. 15th 2022 as early stage infects patients and those infected after Jan. 1st 2023 as late stage infects patients (Figure 1). Information of patients infected within the two periods were then extracted from the background survey and the clinical features of patients in the two groups were then compared. This data generated in this study are properly anonymized to minimize the risk of leaking personal sensitive information. This study is approved by the institutional review board of Xi’an Jiaotong University Health Science Center (No.2023-7).

**Figure 1.**
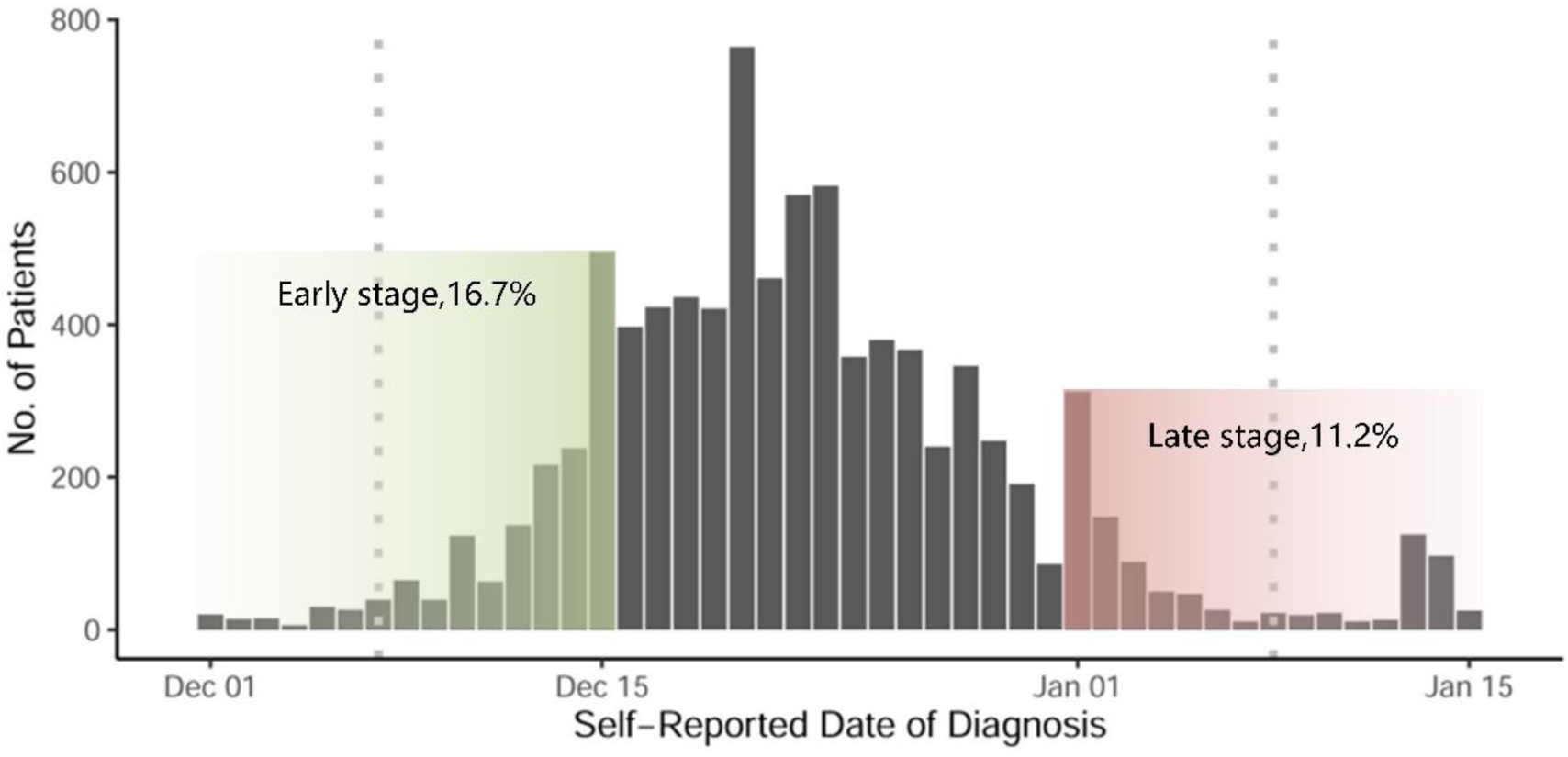
Time distribution of self-reported date of diagnosis for the COVID-19 patients. Green area: early-stage infection (Dec.15st, 2022-Jan.1st2023). Red area: late infection. Left dotted line: The date when the 10 new COVID easing steps were announced (Dec. 7th, 2022). Right dotted line: The official beginning of the Spring Festival travel rush (Jan. 7th, 2023).

### Statistical Analysis

The Student’s t-test and χ^2^ test were utilized for significance tests on continuous and categorical data, respectively. A slope graph was made to visualize the differences of the symptom patterns between COVID-19 patients infected in early and late stages. Multiple logistic regression model was constructed to evaluate the effects of periods of infections on patterns of COVID-19 symptoms while adjusting for some potential confounder. A forest plot was created to summarize the results of multiple logistic regression. Statistical computing software R v4.3.2 and relevant packages were utilized for statistical analysis and results visualization[18].

## Results

### Characteristics of the Study Participants

According to the background survey, a total of 10,258 study participants (out of 12,111 people) were infected with SARS-Cov-2, representing a cumulative infection rate of 84.7% during the COVID-19 pandemic in Shaanxi Province [13]. Information of 2,854 COVID-19 patients, comprising of 1,710 (16.7%) patients infected during the early stage (before Dec. 15th 2022) and 1,144 (11.2%) during the late stage (after Jan. 1st 2023), were extracted from the background survey (Supplementary Figure S1 and Table 1). The average age of infected patients in the early and late stages (40.66 and 44.96) was significantly different (t=-6.27, *p*<0.001). Early stage infects patients included 853 males (49.9%) and 857 females (50.1%), while late stage infects patients were comprised of included 567 males (49.6%) and 577 females (50.4%). Among those who were infected in the early stage, 928 patients (54.3%) resided in urban areas, surpassing the 782 patients (45.7%) in townships. On the other hand, among the late stage infects patients, 651 (56.9%) were from townships, which exceeded the 493 patients (43.1%) from urban areas, indicating a significant distinction between the two groups (χ^2^=33.792, *p* < 0.001). In addition, the average No. of family members of early infected patients was 3.0, which was significantly higher than the No. of family members for those late infected patients, which was 2.6 (t=7.25, *p*<0.001).

**Table 1.** Demographic characteristics of the study participants.

| Variable | Early stage(N=1710) | Late stage(N=624) | Statistics |  |
| --- | --- | --- | --- | --- |
| | | | (t / $\chi^2$) | P-value |
| Age | 40.7±16.7 | 45.0±18.8 | -6.27 | <0.001 |
| Sex (%) |  |  |  |  |
| *male* | 853(49.9) | 567(49.6) |  |  |
| *female* | 857(50.1) | 577(50.4) | 0.02 | 0.9 |
| Residence (%) |  |  |  |  |
| *urban* | 928(54.3) | 493(43.1) | 33.79 | <0.001 |
| *rural* | 782(45.7) | 651(56.9) |  |  |
| No. of family members | 3.0±1.6 | 2.6±1.5 | 7.25 | <0.001 |
| No. of household transmission cases | 2.6±1.7 | 2.5±3.4 | 1.67 | 0.1 |
| No. of pre-existing conditions (%) |  |  |  |  |
| 1 | 300(17.5) | 293(25.6) | 28.47 | <0.001 |
| 2 | 56(3.3) | 37(3.2) |  |  |
| $\geq 3$ | 17(1.0) | 15(1.3) | | |
| *None of basic disease* | 1337(78.2) | 799(69.8) |  |  |
| Basis for diagnosis (%) |  |  |  |  |
| *asymptomatic* | 140(8.2) | 111(9.7) | 32.82 | <0.001 |
| *obvious self-perceived* |  |  |  |  |
| *symptoms* | 988(57.8) | 712(62.2) |  |  |
| *positive antigen test* | 255(14.9) | 193(16.9) |  |  |
| *positive nucleic acid test* | 327(19.1) | 128(11.2) |  |  |

\* The Student's t-test and $\chi^2$ test were utilized for significance tests on continuous and categorical data, respectively.

### Significant differences in patterns of COVID-19 symptoms between early and late stage infects patients

Among the symptoms of infected patients, 1568 (54.94%) had a cough, 1499 (52.52%) had a fever, and 1161 (40.68%) had a sore throat (Table S1). Most of the COVID-19 typical symptoms (11 out of 12) were found to be more frequent in COVID-19 patients infected during the early stage compared to those infected during late stage (Table 2 and Figure 2). Nine of them showed statistical significance including fever (χ^2^=93.42, *P*<0.001), cough (χ^2^=8.97, *P*=0.003), muscle or body pain (χ^2^=17.57, *P*<0.001), headache (χ^2^=8.28, *P*=0.004), sore throat (χ^2^=10.10, *P=*0.002), nausea or vomiting (χ^2^=8.40, *P*=0.004), loss of taste or smell(χ^2^=45.05, *P*<0.001) and chills(χ^2^=8.17, *P*=0.004), were identified significantly Only one symptom, runny nose (χ^2^=12.56, *P*<0.001), was found in significantly more patients infected during the late stage of the pandemic compared to early stage.

**Figure 2.**
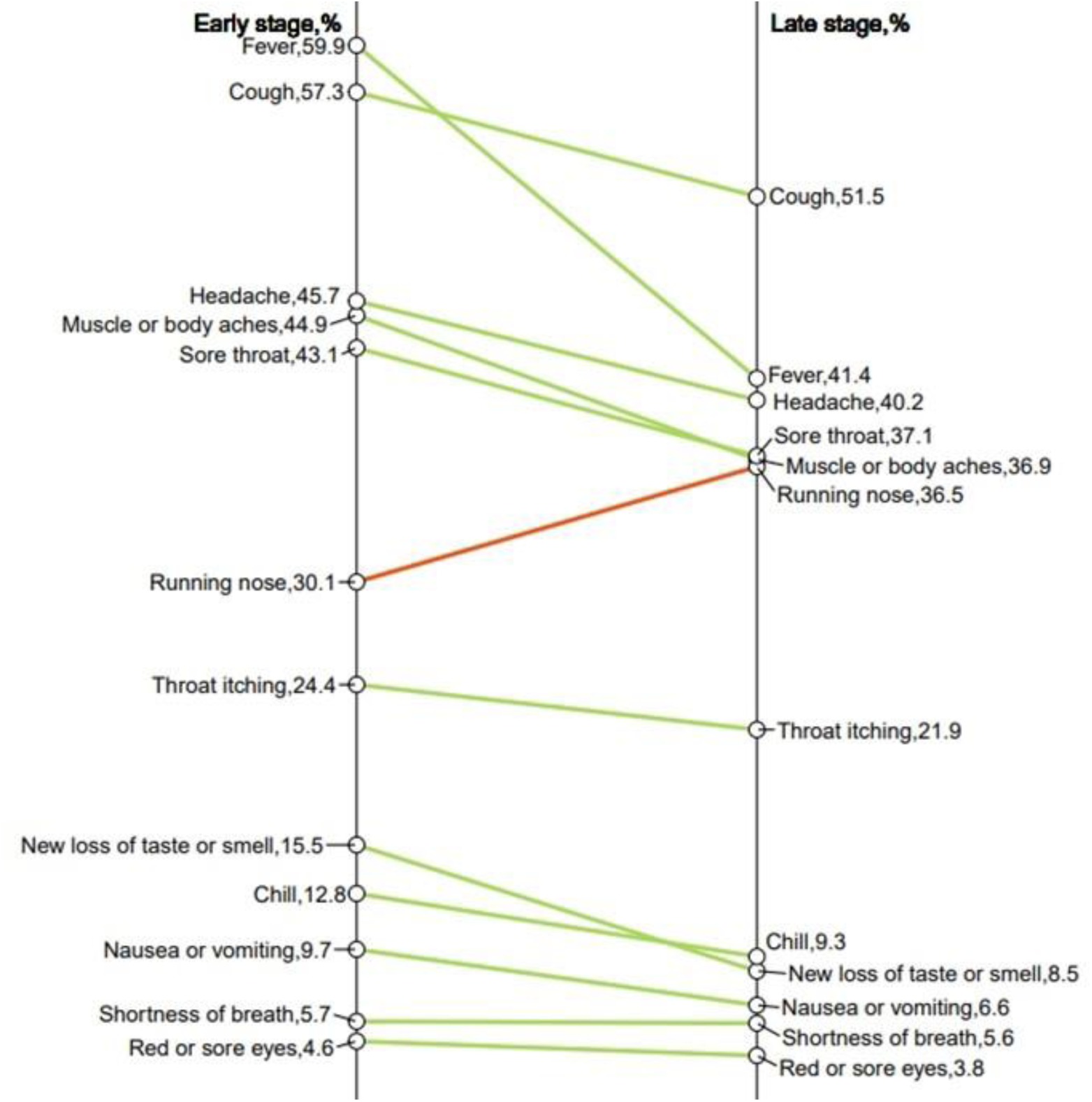
Differences in proportions of the COVID-19 typical symptoms between early and late infected persons. Left vertical line: early-stage. Right line: late-stage.

**Table 2.**
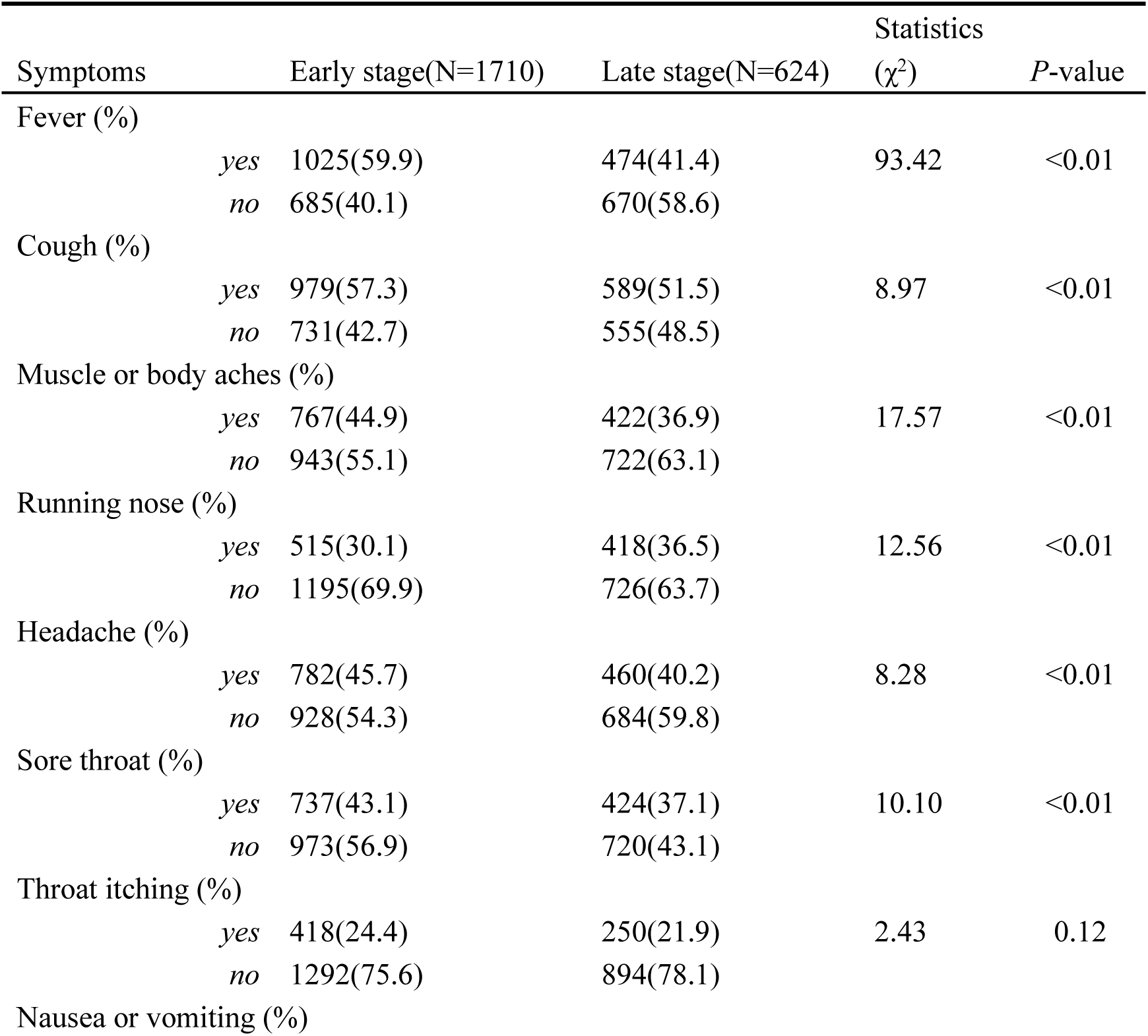
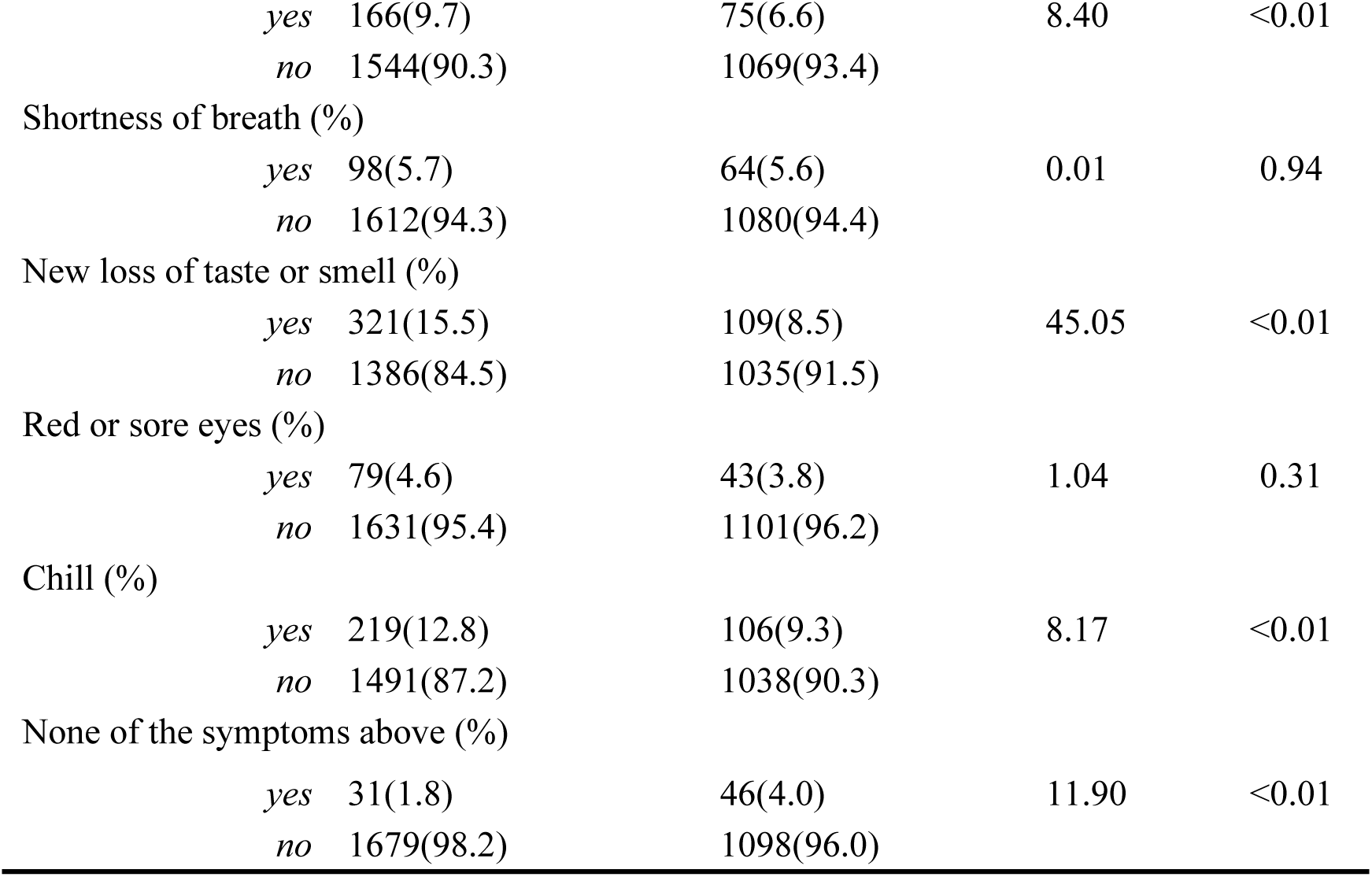
Comparison of early and late symptoms in COVID-19 infected persons.

To further explore whether the significant differences in symptom patterns between COVID-19 patients infected during the early and late stages were influenced by other related factors, multiple logistic regression models were constructed for each symptom (Coding scheme could be identified in supplementary Table S14). Age, sex, residence, pre-existing conditions and basis of diagnosis were included in each model as covariates. Among the 12 major symptoms investigated in the background survey, significance of 9 symptoms were kept after being adjusted for multiple variables (Figure 3). Infected during late stage of the COVID-19 pandemic is significantly associated with decreased frequency of fever (OR [95%CI] =0.52[0.45-0.62]), cough (OR [95%CI] =0.82[0.70-0.96]), muscle or body aches(OR [95%CI] =0.76[0.64-0.89]), headache (OR [95%CI] =0.83[0.71-0.98]), sore throat (OR [95%CI] =0.77[0.66-0.90]), nausea or vomiting (OR [95%CI] =0.70[0.52-0.94]), loss of taste or smell(OR [95%CI] =0.47[0.37-0.60]), chill (OR [95%CI] =0.74[0.57-0.95]). On the other hand, infected during late stage is significantly associated with increased frequency of runny nose (OR [95%CI] =1.29[1.09-1.52]). Full results of the multiple logistic regression analysis were summarized in Supplementary Table S2-S13.

**Figure 3.**
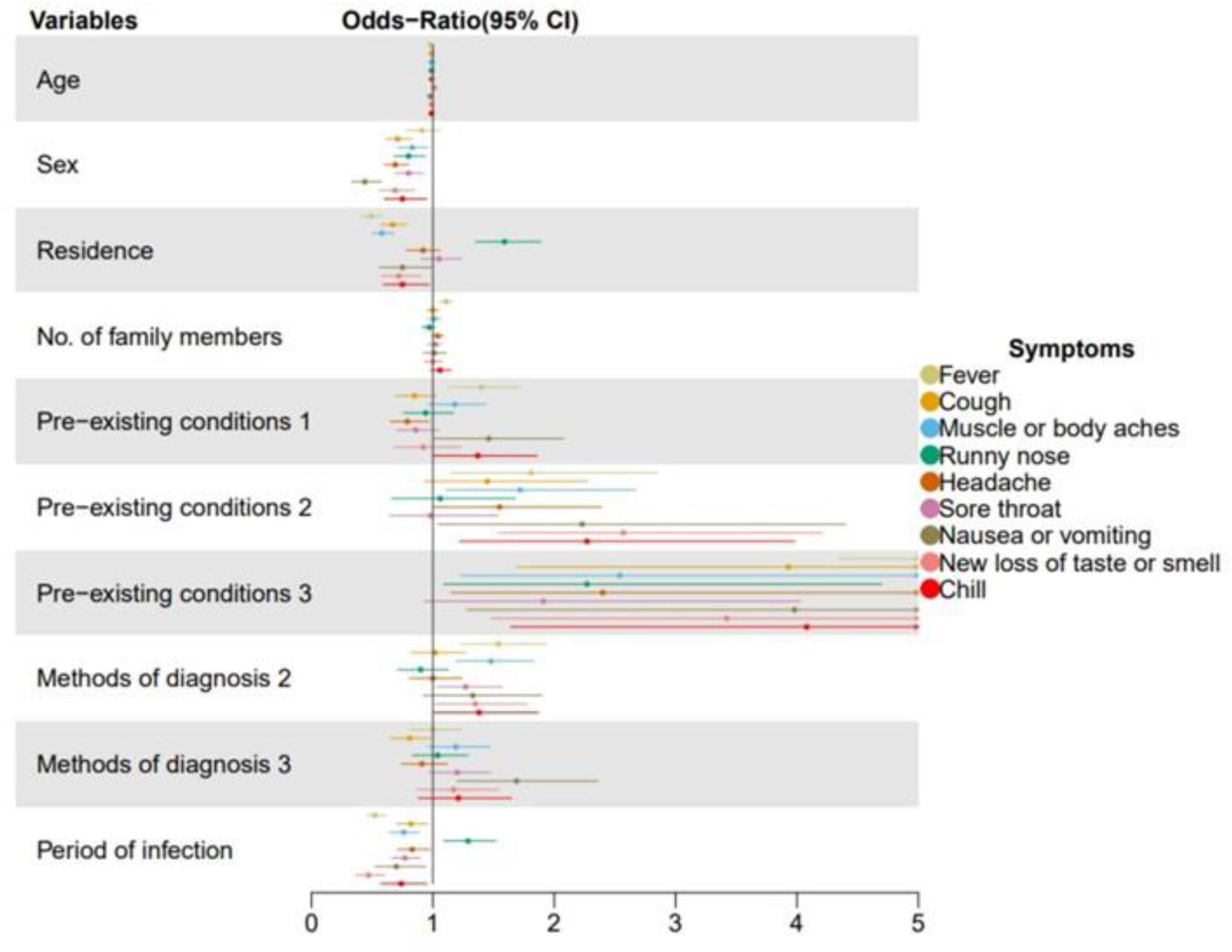
Forest map of multiple regression results with symptom outcomes. Variable: Independent variable. Symptoms: Outcome variable. Each color represents a multiple regression result for one symptom.

## Discussion

In the present study, significant higher proportions were identified for multiple symptoms of COVID-19 including fever, cough, headache, muscle or body aches, score throat, nausea or vomiting, loss of taste or smell, and chill in patients during the early stage comparing to the late stage of the outbreak. This pattern was kept after being adjusted for multiple factors. To the best of our knowledge, the current study is the first one to report the symptom features of COVID-19 among individuals infected during different stages of a single outbreak, although some previous studies have compared the symptom features of COVID-19 among individuals infected during different outbreaks caused by different SARS-Cov-2 variants.

Besides periods of infections during the outbreak, several other factors including age, sex, number of family members, pre-existing conditions, and the methods of diagnosis were also identified to be significantly associated with the proportions of symptoms. A previous study suggested that men might be more susceptible to COVID-19 due to higher concentrations of angiotensin-converting enzyme 2 (ACE2) in males [19]. Similar results were also reported in a study comparing the characteristics of infection in Delta and Omicron in southern Sweden [20]. However, the impact of gender on COVID-19 symptoms remains largely unknown. In this sense, the current study has provided additional details for those previous work. Female are more likely than male to develop certain symptoms in the current study despite that female sex appears to be a protective factor for infection of SARS-Cov-2. Patients with pre-existing conditions were more likely to have certain symptoms in the current study. Bottari *et al*. has reported that adults with combined diabetes and cardiovascular disease were more likely to have severe COVID-19 symptoms and higher mortality[21].

The associations between infection periods and symptom patterns could have three possible explanations. The first one is that the patients at early and late stages of the outbreak were infected by different SARS-CoV-2 variants. However, in the context of a rapidly escalating short-term outbreak of COVID-19, typically only one dominant SARS-Cov-2 variant is present. Another potential explanation is that late-stage COVID-19-infected individuals might have better preparedness for relevant medications, resulting in a lower proportion of occurrence for associated symptoms among these patients compared to those who infected earlier. However, this might not be true. After the onset of the COVID-19 pandemic in December 2022, major cities in China experienced a severe medicine shortage [22]. Many COVID-19-related medications, such as painkillers or antipyretics, were nearly impossible to purchase despite that the relevant authorities in China mainland had introduced policies and measures such as increasing the production and precise delivery of relevant drugs [23–24]. This implies that, compared to the patients infected at early stage, patients infected at late stage may not necessarily have better preparedness in terms of relevant medications. This trend can be validated by the data from background survey of this current study. In the comparison of drug sources between early and late infected persons, it can be found that the preparation of early infected persons is significantly higher than that of late infected persons in terms of antigen detection reagents and cough medicines (Table S15). In addition, the sources of medication for both early and late infected patients were mainly daily household preparations and self-purchase, but a higher percentage of late infected patients obtained their medication through official or unofficial mutual aid channels than in the early stages. The last hypothesis on the differences in symptoms between patients infected at early and late stages is about the psychological status of the patients. Before the COVID-19 pandemics of Dec. 2022, China adhered to a long-term “dynamic zero-COVID” policy for COVID-19 prevention and control[9]. As a result, the country has not experienced significant nationwide outbreaks of COVID-19. Given that much of the information about COVID-19 available to residents comes from the internet and media coverage, their perceptions of the omicron variant may be influenced by biases. A study examining the relationship between COVID-19 and anxiety, as well as other associated emotions[25], revealed that a significant source of anxiety among the majority of adults is the concern surrounding COVID-19 and its related illnesses.

Furthermore, children also express apprehensions regarding conflicts and safety challenges stemming from the COVID-19 situation. In addition, a study conducted by Dou. *et al*., it was found that during the COVID-19 pandemic, different groups of people suffered from psychological distress and lower levels of somatization symptoms to varying degrees[26]. Conversely, individuals who were infected at a later stage appear to possess a greater array of strategies for managing COVID-19, along with a more positive outlook. This is attributed to their exposure to information about acquaintances and friends who have experienced infection, as well as media coverage. Consequently, these late infects individuals are less prone to experiencing emotional challenges like anxiety and worry.

There are some limitations in this study. firstly, we did not test the SARS-Cov-2 variants of our study participants, which made it difficult for us to validate potential effects of the virus variants on the differences in symptoms among COVID-19 infected individuals. Another limitation is that, as an observational study, this study used self-completed questionnaires by the study participants to record their symptoms, and there were no objective medical records to corroborate them, which may result in bias.

The findings from this study indicate that individuals infected with the Omicron variant at an earlier stage of a COVID-19 outbreak are more prone to developing certain symptoms compared to those infected at late stage. The outcomes of this study offer valuable insights for future pandemic prevention and control measures while follow-up surveys are still needed to further validate the current results.

## Data Availability

The data that support the findings of this study are available on request from the corresponding author TZ. The data are not publicly available due to local laws, regulations and restrictions.

## Acknowledgements

We want to thank the student research volunteers for their focusing on work during winter vacations. We also want to thank the study participants. Without them no meaningful research could be done.

## Provenance and peer review

Not commissioned; externally peer reviewed.

## Ethics approval and consent to participate

All participants received informed consent. This study was approved by the Ethics Review Committee of Xi ’an Jiaotong University Health Science Center (No. 2023-7). All participants fully read the informed consent form and clicked to confirm before completing the questionnaire, minors were accompanied by their parents to complete the questionnaire.

## Consent for publication

Not applicable.

## Authors Contributions

SZ proposed the first idea of performing this study. CY, TZ, QW, SZ, YZ, YW, and GZ then thoroughly discussed the idea and make it a completed study design. BM directed the large scale field work. YN, SC, JL, HH, ZW, and LL contributed significantly in the field work and data collection. CY, TZ, PA, YY and MS performed data analyses. CY and TZ drafted the first version of manuscript. All the authors contributed to the revisions of the manuscript. All authors have read and approved the manuscript.

