## Supplementary Materials for "Systematic differences were identified for symptoms of Omicron infected individuals between early and late stage of the COVID-19 pandemic in China"

**Supplementary Tables**

| **Supplementary Table S1.**Proportions of typical COVID-19 symptoms reported by the patients. | | |
| --- | --- | --- |
| Symptoms | No. of Patients | Proportion in the Patients, % |
| Fever | 1499 | 52.52 |
| Cough | 1568 | 54.94 |
| Headache | 1242 | 43.52 |
| Muscle or body aches | 1189 | 41.66 |
| Runny nose | 933 | 32.96 |
| Sore throat | 1161 | 40.68 |
| Throat itching | 668 | 23.41 |
| New loss of taste or smell | 430 | 15.07 |
| Chill | 325 | 11.39 |
| Nausea or vomiting | 241 | 8.44 |
| Shortness of breath | 162 | 5.68 |
| Red or sore eyes | 122 | 4.27 |
| None of the symptoms above | 77 | 2.70 |

**Supplementary Table S2.** Analysis of the results of the multiple regression model

(Symptom 1: Fever)

| Variable | OR1 | LL1 | UL1 | *P*-value1 |
| --- | --- | --- | --- | --- |
| Age | 0.98 | 0.97 | 0.98 | <0.001 |
| Sex | 0.91 | 0.78 | 1.06 | 0.242 |
| Residence | 0.49 | 0.42 | 0.58 | <0.001 |
| No. of family members | 1.11 | 1.05 | 1.17 | <0.001 |
| Pre-existing conditions 1 | 1.40 | 1.13 | 1.72 | 0.002 |
| Pre-existing conditions 2 | 1.81 | 1.15 | 2.85 | 0.010 |
| Pre-existing conditions 3 | 10.85 | 4.35 | 33.03 | <0.001 |
| Methods of diagnosis 2 | 1.54 | 1.23 | 1.94 | <0.001 |
| Methods of diagnosis 3 | 1.00 | 0.81 | 1.24 | 0.989 |
| period of infection | 0.52 | 0.45 | 0.62 | <0.001 |

**Supplementary Table S3.** Analysis of the results of the multiple regression model

(Symptom 2: Cough)

| Variable | OR2 | LL2 | UL2 | *P*-value2 |
| --- | --- | --- | --- | --- |
| Age | 0.99 | 0.99 | 1.00 | 0.382 |
| Sex | 0.71 | 0.61 | 0.83 | <0.001 |
| Residence | 0.67 | 0.57 | 0.79 | <0.001 |
| No. of family members | 1.00 | 0.95 | 1.05 | 0.983 |
| Pre-existing conditions 1 | 0.85 | 0.69 | 1.03 | 0.098 |
| Pre-existing conditions 2 | 1.45 | 0.93 | 2.28 | 0.106 |
| Pre-existing conditions 3 | 3.93 | 1.69 | 10.73 | 0.003 |
| Methods of diagnosis 2 | 1.02 | 0.82 | 1.27 | 0.856 |
| Methods of diagnosis 3 | 0.81 | 0.65 | 0.99 | 0.044 |
| period of infection | 0.82 | 0.70 | 0.96 | 0.015 |

**Supplementary Table S4**. Analysis of the results of the multiple regression model

(Symptom 3: Muscle or body aches)

| Variable | OR3 | LL3 | UL3 | *P*-value3 |
| --- | --- | --- | --- | --- |
| Age | 0.99 | 0.99 | 1.00 | 0.443 |
| Sex | 0.83 | 0.71 | 0.96 | 0.015 |
| Residence | 0.58 | 0.50 | 0.68 | <0.001 |
| No. of family members | 1.01 | 0.96 | 1.07 | 0.638 |
| Pre-existing conditions 1 | 1.18 | 0.96 | 1.44 | 0.117 |
| Pre-existing conditions 2 | 1.72 | 1.11 | 2.67 | 0.015 |
| Pre-existing conditions 3 | 2.54 | 1.23 | 5.41 | 0.013 |
| Methods of diagnosis 2 | 1.48 | 1.19 | 1.83 | <0.001 |
| Methods of diagnosis 3 | 1.19 | 0.96 | 1.47 | 0.107 |
| period of infection | 0.76 | 0.64 | 0.89 | <0.001 |

**Supplementary Table S5.** Analysis of the results of the multiple regression model

(Symptom 4: Runny nose)

| Variable | OR4 | LL4 | UL4 | *P*-value4 |
| --- | --- | --- | --- | --- |
| Age | 0.99 | 0.99 | 1.00 | 0.077 |
| Sex | 0.80 | 0.68 | 0.94 | 0.006 |
| Residence | 1.59 | 1.35 | 1.89 | <0.001 |
| No. of family members | 0.97 | 0.91 | 1.02 | 0.218 |
| Pre-existing conditions 1 | 0.94 | 0.76 | 1.17 | 0.592 |
| Pre-existing conditions 2 | 1.06 | 0.66 | 1.68 | 0.797 |
| Pre-existing conditions 3 | 2.27 | 1.09 | 4.70 | 0.026 |
| Methods of diagnosis 2 | 0.90 | 0.71 | 1.13 | 0.378 |
| Methods of diagnosis 3 | 1.04 | 0.83 | 1.29 | 0.736 |
| period of infection | 1.29 | 1.09 | 1.52 | 0.002 |

**Supplementary Table S6.** Analysis of the results of the multiple regression model

(Symptom 5: Headache)

| Variable | OR5 | LL5 | UL5 | *P*-value5 |
| --- | --- | --- | --- | --- |
| Age | 0.99 | 0.99 | 1.00 | 0.141 |
| Sex | 0.69 | 0.60 | 0.80 | <0.001 |
| Residence | 0.92 | 0.78 | 1.06 | 0.289 |
| No. of family members | 1.04 | 0.99 | 1.09 | 0.146 |
| Pre-existing conditions 1 | 0.79 | 0.65 | 0.97 | 0.026 |
| Pre-existing conditions 2 | 1.55 | 1.00 | 2.39 | 0.047 |
| Pre-existing conditions 3 | 2.40 | 1.15 | 5.10 | 0.022 |
| Methods of diagnosis 2 | 1.00 | 0.81 | 1.24 | 0.994 |
| Methods of diagnosis 3 | 0.91 | 0.74 | 1.12 | 0.371 |
| period of infection | 0.83 | 0.71 | 0.98 | 0.023 |

**Supplementary Table S7.** Analysis of the results of the multiple regression model

(Symptom 6: Sore throat)

| Variable | OR6 | LL6 | UL6 | *P*-value6 |
| --- | --- | --- | --- | --- |
| Age | 1.01 | 1.00 | 1.01 | 0.013 |
| Sex | 0.80 | 0.69 | 0.93 | 0.004 |
| Residence | 1.05 | 0.90 | 1.24 | 0.516 |
| No. of family members | 1.02 | 0.96 | 1.07 | 0.563 |
| Pre-existing conditions 1 | 0.86 | 0.70 | 1.05 | 0.132 |
| Pre-existing conditions 2 | 0.98 | 0.64 | 1.54 | 0.99 |
| Pre-existing conditions 3 | 1.91 | 0.93 | 4.03 | 0.082 |
| Methods of diagnosis 2 | 1.27 | 1.03 | 1.57 | 0.027 |
| Methods of diagnosis 3 | 1.20 | 0.97 | 1.48 | 0.083 |
| period of infection | 0.77 | 0.66 | 0.90 | 0.001 |

**Supplementary Table S8.** Analysis of the results of the multiple regression model

(Symptom 7: Throat itching)

| Variable | OR7 | LL7 | UL7 | *P*-value7 |
| --- | --- | --- | --- | --- |
| Age | 0.99 | 0.99 | 1.00 | 0.221 |
| Sex | 0.76 | 0.64 | 0.97 | 0.002 |
| Residence | 1.10 | 0.91 | 1.32 | 0.319 |
| No. of family members | 1.08 | 1.02 | 1.14 | 0.013 |
| Pre-existing conditions 1 | 0.89 | 0.70 | 1.13 | 0.335 |
| Pre-existing conditions 2 | 1.58 | 0.97 | 2.53 | 0.060 |
| Pre-existing conditions 3 | 2.88 | 1.37 | 5.96 | 0.005 |
| Methods of diagnosis 2 | 1.21 | 0.94 | 1.55 | 0.137 |
| Methods of diagnosis 3 | 1.74 | 1.38 | 2.18 | <0.001 |
| period of infection | 0.94 | 0.78 | 1.13 | 0.502 |

**Supplementary Table S9.** Analysis of the results of the multiple regression model

(Symptom 8: Nausea or vomiting)

| Variable | OR8 | LL8 | UL8 | *P*-value8 |
| --- | --- | --- | --- | --- |
| Age | 0.98 | 0.97 | 0.99 | <0.001 |
| Sex | 0.44 | 0.33 | 0.58 | <0.001 |
| Residence | 0.75 | 0.56 | 0.99 | 0.046 |
| No. of family members | 1.01 | 0.92 | 1.11 | 0.832 |
| Pre-existing conditions 1 | 1.46 | 1.01 | 2.08 | 0.040 |
| Pre-existing conditions 2 | 2.23 | 1.04 | 4.40 | 0.027 |
| Pre-existing conditions 3 | 3.98 | 1.28 | 10.26 | 0.007 |
| Methods of diagnosis 2 | 1.33 | 0.92 | 1.90 | 0.119 |
| Methods of diagnosis 3 | 1.69 | 1.20 | 2.36 | 0.002 |
| period of infection | 0.70 | 0.52 | 0.94 | 0.019 |

**Supplementary Table S10.** Analysis of the results of the multiple regression model

(Symptom 9: Shortness of breath)

| Variable | OR9 | LL9 | UL9 | *P*-value9 |
| --- | --- | --- | --- | --- |
| Age | 0.99 | 0.98 | 1.00 | 0.157 |
| Sex | 0.49 | 0.35 | 0.69 | <0.001 |
| Residence | 0.62 | 0.47 | 0.88 | 0.008 |
| No. of family members | 1.03 | 0.91 | 1.15 | 0.658 |
| Pre-existing conditions 1 | 2.43 | 1.61 | 3.67 | <0.001 |
| Pre-existing conditions 2 | 4.44 | 2.13 | 3.67 | <0.001 |
| Pre-existing conditions 3 | 18.90 | 7.91 | 43.54 | <0.001 |
| Methods of diagnosis 2 | 1.89 | 1.25 | 2.83 | 0.002 |
| Methods of diagnosis 3 | 1.57 | 1.00 | 2.39 | 0.043 |
| period of infection | 0.96 | 0.68 | 1.35 | 0.831 |

**Supplementary Table S11.** Analysis of the results of the multiple regression model

(Symptom 10: New loss of taste or smell)

| Variable | OR10 | LL10 | UL10 | *P*-value10 |
| --- | --- | --- | --- | --- |
| Age | 0.99 | 0.989 | 1.00 | 0.288 |
| Sex | 0.69 | 0.56 | 0.85 | <0.001 |
| Residence | 0.72 | 0.58 | 0.90 | 0.004 |
| No. of family members | 1.00 | 0.93 | 1.08 | 0.932 |
| Pre-existing conditions 1 | 0.92 | 0.68 | 1.23 | 0.563 |
| Pre-existing conditions 2 | 2.57 | 1.54 | 4.21 | <0.001 |
| Pre-existing conditions 3 | 3.42 | 1.48 | 7.45 | 0.003 |
| Methods of diagnosis 2 | 1.35 | 1.02 | 1.78 | 0.034 |
| Methods of diagnosis 3 | 1.17 | 0.87 | 1.54 | 0.285 |
| period of infection | 0.47 | 0.37 | 0.60 | <0.001 |

**Supplementary Table S12.** Analysis of the results of the multiple regression model

(Symptom 11: Red or sore eyes)

| Variable | OR11 | LL11 | UL11 | *P*-value11 |
| --- | --- | --- | --- | --- |
| Age | 0.99 | 0.98 | 1.00 | 0.072 |
| Sex | 0.75 | 0.6 | 0.95 | 0.055 |
| Residence | 0.75 | 0.59 | 0.97 | 0.379 |
| No. of family members | 1.06 | 0.98 | 1.15 | 0.857 |
| Pre-existing conditions 1 | 1.37 | 1.00 | 1.86 | 0.928 |
| Pre-existing conditions 2 | 2.27 | 1.22 | 3.98 | 0.118 |
| Pre-existing conditions 3 | 4.08 | 1.64 | 9.22 | 0.006 |
| Methods of diagnosis 2 | 1.38 | 1.00 | 1.87 | 0.134 |
| Methods of diagnosis 3 | 1.21 | 0.88 | 1.65 | 0.788 |
| period of infection | 0.74 | 0.57 | 0.95 | 0.383 |

**Supplementary Table S13.** Analysis of the results of the multiple regression model

(Symptom 12: Chill)

| Variable | OR12 | LL12 | UL12 | *P*-value12 |
| --- | --- | --- | --- | --- |
| Age | 0.99 | 0.98 | 1.00 | 0.008 |
| Sex | 0.75 | 0.60 | 0.95 | 0.019 |
| Residence | 0.75 | 0.59 | 0.97 | 0.026 |
| No. of family members | 1.06 | 0.98 | 1.15 | 0.156 |
| Pre-existing conditions 1 | 1.37 | 1.00 | 1.86 | 0.05 |
| Pre-existing conditions 2 | 2.27 | 1.22 | 3.98 | 0.006 |
| Pre-existing conditions 3 | 4.08 | 1.64 | 9.22 | 0.001 |
| Methods of diagnosis 2 | 1.38 | 1.00 | 1.87 | 0.043 |
| Methods of diagnosis 3 | 1.21 | 0.88 | 1.65 | 0.236 |
| period of infection | 0.74 | 0.57 | 0.95 | 0.018 |

**Supplementary Table S14.** Coding scheme for multiple logistic regression

| Factor | |  | variable | Coding scheme |
| --- | --- | --- | --- | --- |
| Age | |  | X1 | unit: year |
| Sex | |  | X2 | male=1, female=0 |
| Residence | |  | X3 | urban=1, rural=2 |
| No. of family members | |  | X4 | - |
| Pre-existing conditions | |  | X5 | none of basic disease=0, suffers from 2 disease=1, suffers from 1 disease=2, suffering from 3 or more diseases=3 |
| Methods of diagnosis | |  | X6 | self-perceived symptom=1, positive antigen test=2, positive nucleic acid test=3 |
| Period of infection | |  | X7 | early stage=1, late stage=2 |
| Symptoms | |  | Y | yes=1, no=0 |
|  | *Using the term with a categorical variable of 0 as the comparison term in the multiple regression model | | | |

**Supplementary Table S15.** Drug readiness for early and late infected persons

| Variables | Early Stage | Late Stage | χ^2^ | *P*-value |
| --- | --- | --- | --- | --- |
| Antigen Detection Reagents | |  |  |  |
| *yes* | 356(20.8) | 199(17.4) |  |  |
| *no* | 1354(79.2) | 945(82.6) | 4.91 | <0.001 |
| Antifebrile |  |  |  |  |
| *yes* | 1222(71.5) | 825(72.1) |  |  |
| *no* | 488(28.5) | 319(27.9) | 0.11 | 0.736 |
| Antitussive |  |  |  |  |
| *yes* | 1051(61.5) | 644(56.3) |  |  |
| *no* | 659(38.5) | 500(43.7) | 7.38 | <0.001 |
| Oximetry equipment |  |  |  |  |
| *yes* | 60(3.5) | 46(4.0) | 0.37 | 0.543 |
| *no* | 1650(96.5) | 1098(96.0) |  |  |
| Ventilator |  |  |  |  |
| *yes* | 4(0.2) | 4(0.3) |  |  |
| *no* | 1706(99.8) | 1140(99.7) | 0.04 | 0.832 |
| Oxygen Generator |  |  |  |  |
| *yes* | 28(1.6) | 20(1.7) |  |  |
| *no* | 1682(98.4) | 1124(98.3) | 0.01 | 0.939 |
| Other Medicines |  |  |  |  |
| *yes* | 31(1.8) | 18(1.6) |  |  |
| *no* | 1679(98.2) | 1126(98.4) | 0.11 | 0.737 |
| Sources of Drugs |  |  |  |  |
| *daily preparation of the household* | 625(44.9) | 403(43.4) |  |  |
| *personal purchase* | 750(53.9) | 508(54.7) |  |  |
| *official or unofficial mutual aid channels* | 13(0.9) | 14(1.5) |  |  |
| *other sources* | 3(0.2) | 4(0.4) | 2.78 | 0.426 |

**Supplementary Figures**

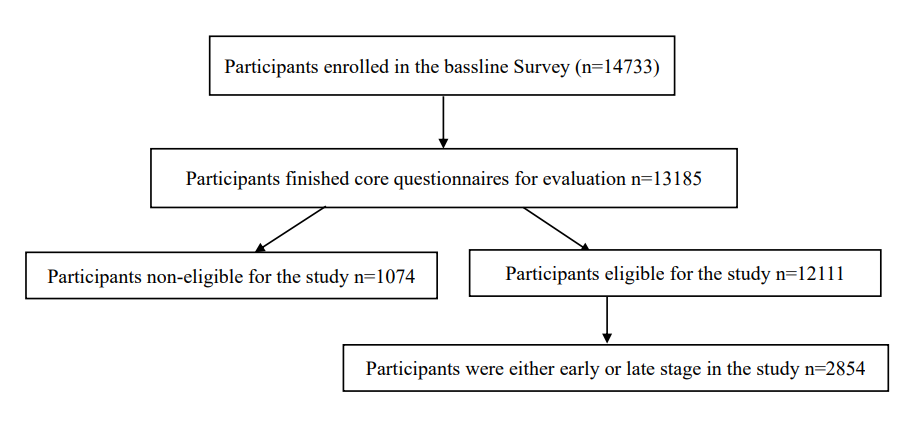

**Supplementary Figure S1.** Flow chart of the study participants enrollment.

**A sample of the survey questionnaire**

**COVID-19 Surveillance in Urban And Rural Areas Of Shaanxi Province (Baseline Survey)**

Record ID：

**1.** Mobile phone number:

**2.** Name:

**3.** ID number:

**4.** Current age (years):

**5.** Gender:

**6.** Current residence:

**7.** What is your type of residence?

 Urban area

 Villages and towns

**8.** What is your usual residence?

 This village/community

 Outside village/outside community in this county

 Counties outside the city

 City outside the province of shaanxi

 Provincial

 Overseas

**9.** How many people live in your family (besides you)?

**10.** How many people in your family are currently infected with COVID-19 (besides you)?

**11.** Are you the head of the household?

 Yes

 No

**11.1** Name of householder:

**12.** Do you have any of the following underlying medical conditions (select at least 1)?

 Hypertension

 Diabetes

 Chronic Obstructive Pulmonary Disease

 Chronic Bronchitis

 Asthma

 CVD(Cardiovascular Disease)

 Stroke

 Chronic Nephrosis

 Chronic Liver Disease

 Hematological System Diseases

 Disease Of Immune System

 Malignant Tumor

 Other Underlying Diseases

 None Of The Above

**13.** Are you infected with COVID-19?

 Never infected

 Infected with once

 Infected with two or more

**14.** What was your basis for confirming the infection?

 ConsciouslyAsymptomatic

 Self-evident symptom

 Antigen test positive

 Nucleic acid test positive

**14.1** The date of your antigen positive test is:

**14.2** The date of your positive nucleic acid test is:

**15.** During infection, do you have the following symptoms?

 Body temperature over 38.0℃:

 Cough (if you have a basis for coughing, more severe than usual)

 Muscle pain (if you usually have muscle pain, is it worse?)

 Snotty

 [Headache](javascript:;)

 Sore throat

 Itchy throat

 Nausea, vomiting, or diarrhea

 Polypnea

 Unable to taste or smell

 Red or sore eyes

 Shiver

 None of the above

16. What is the degree of pain all over?

 Mild pain

The pain is moderate and tolerable

 Unbearable pain

17. Have you sought medical attention after infection?

 Outpatient/emergency

 In hospital

 Not seen a doctor

18. Do you still have symptoms?

 Yes, it's still serious

 Yes, but it's getting better

 No

 19. Have you been tested for antigens in the last 48 hours?

 Yes, negative

 Yes, positive

 No

20. Have you taken a nucleic acid test in the last 48 hours?

 Yes, negative

 Yes, positive

 No

21. In the past 48 hours, have you participated in any gathering activities (five or more people in a confined indoor environment)?

 Yes

 No

22. How many people have you had close contact with in the last 48 hours?

23. What medical drugs do you have at present?

 Antigen detection reagent

 Febrifuge

 Pectoral

 Oximetry equipment (e.g. Clip-on oximeter)

 Breathing machine

 Oxygenerator

 Other

 None of the above

24. What is the source of all your medical drugs?

 Household supplies

 Purchase by oneself

 Official or unofficial channels for mutual assistance

 Other

25. What is your preferred means of transportation at present?

 Walk

 Bicycle

 Electromobile

 Motorbike

 Subway

 Bus

 Private car

 Other

26. Do you have any plans to visit relatives across provinces/cities during the Spring Festival?

 Yes, interprovincial

 Yes, within the province and across the city

 Yes, the city is cross-county

 Yes, county flow

 No

**First follow-up**

Record ID：

1. Have any relatives returned since the last survey?

 Yes

 No

2. Have you been infected with COVID-19 since the last survey?

 Uninfected

 Infected

3. Do you still have symptoms?

 Yes, it's still serious

 Yes, but it's getting better

 No

4. Have you been tested for antigens since the last survey?

 Yes, negative

 Yes, positive

 No

5. Have you taken a nucleic acid test since the last investigation?

 Yes, negative

 Yes, positive

 No

6. Since the last survey, have you participated in any gathering activities (more than 5 people in a confined indoor environment)?

 Yes

 No

7. How many people have you had close contact with since your last survey?

8. What medical drugs do you have at present?

 Antigen detection reagent

 Febrifuge

 Pectoral

 Oximetry equipment (e.g. Clip-on oximeter)

 Breathing machine

 Oxygenerator

 Other

 None of the above

9. What is the source of all your medical drugs?

 Household supplies

 Purchase by oneself

 Official or unofficial channels for mutual assistance

 Other
